# Understanding the implementation of antimicrobial resistance policies in Vietnam: a multilayer analysis of the veterinary drug value chain

**DOI:** 10.1101/2024.06.27.24309573

**Authors:** Chloé Bâtie, Nguyen Van Duy, Nguyen Thi Minh Khue, Marisa Peyre, Marion Bordier, Nguyen Thi Dien, Vu Dinh Ton, Flavie Goutard

## Abstract

Reducing antibiotic use in livestock production has been a target for national action plans worldwide. The Vietnamese livestock plan issued in 2017 has, among other objectives, strengthened the regulatory framework for antibiotic use. While a progressive ban on prophylactic antibiotics in feed and the introduction of mandatory prescriptions have been introduced, the level of implementation of these measures remains unknown. This study explores the level of understanding, acceptance, and application of these regulations among veterinary drug value chain stakeholders. An iterative stakeholder mapping and analysis of the veterinary drug value chain was conducted in north and south Vietnam. We organized one focus group discussion in Hanoi with 12 participants and conducted 39 semi-structured interviews with governmental authorities, national research centers, foreign partners, and private stakeholders along the value chain. The discourses were analyzed to (1) map the veterinary drug value chain and interactions among stakeholders, (2) analyze stakeholder’s technical and social capital regarding regulations, and (3) identify factors influencing their implementation. From the map of the veterinary drug value chain, we identified 30 categories of stakeholders. Based on the map, the capital, and the analysis of the discourse, we identified 10 factors that could influence their implementation. These factors included stakeholder perception of the new regulations, their level of knowledge, the availability of technical guidance, conflict of economic interest between stakeholders, scale-related management discrepancies, the technical and financial barriers to the implementation of the regulations at the local level, the presence of an informal distribution channel, international influence, consumer demand for food safety, and the willingness to reduce the burden of antibiotic resistance. It was clearly identified that new regulations are a necessary step to reducing antibiotic usage in Vietnam, but that the lack of local stakeholder involvement combined with technical constraints were barriers to their implementation. The study underlined the need for greater involvement of local stakeholders in the development of regulations as well as the need to mainstream innovations developed by small producers.

## Introduction

The rise in antimicrobial resistance (AMR) threatens to cause a significant number of unnecessary deaths (O’Neill 2016; Naghavi et al. 2024). To address this growing threat, policies have targeted antibiotic usage (ABU) reduction in the human, animal and environmental sectors (Chua et al. 2021). Indeed, AMR is a complex issue that must be addressed from a One Health perspective (McEwen and Collignon 2018). In this regard, the Tripartite Organization (World Health Organization, World Organization for Animal Health, and Food and Agriculture Organization) has established a Global Action Plan calling on member states to take action (OMS 2015). In this context, countries have developed and adopted their own National Action Plan (NAP) for the human and agricultural sectors. Vietnam was the first Southeast Asian country to adopt a national plan, followed by the Philippines in 2015 (Chua et al. 2021). It was approved in 2013, and followed in 2017 by the NAP for livestock and aquaculture (MARD 2017). The main objective was to “tackle the human health risk related to ABR by better controlling the usage of antibiotics in livestock and aquaculture” (MARD 2017). A new NAP with similar objectives was adopted in 2021 (MARD 2021) based on five priorities: raising awareness of antibiotic resistance (ABR), promoting good farming practices and prudent ABU, monitoring ABU and ABR, strengthening international and cross-sectoral collaboration around ABR control, and lastly - the focus of our study - reviewing and enforcing regulations for the improved management of ABU.

Several actions were taken to achieve the latter objective. The Law on Veterinary Medicine (National Assembly 2015) was amended with additional regulations to improve legislation governing ABU management. Antimicrobial growth promoters, which were widely used by Vietnamese farmers (Cuong et al. 2021; Kim et al. 2013) and contribute to antibiotic resistance (McEwen and Collignon 2018), were banned by the Animal Husbandry Law in 2018 (National Assembly 2018). The following decree (n°13/2020/ND-CP), was enacted to gradually cease the use of antibiotics in the feed for prophylaxis (Government of the Socialist Republic of Vietnam 2020). A roadmap of implementation from 2020 to 2025 was issued, starting with high-priority critically important antibiotics, followed by the other antibiotics on the WHO list of critical classes, and finally for all antibiotic classes (WHO 2019) with a complete ban planned for 2025. In 2020, a circular (n°122020TT-BNNPTNT) was issued to establish the prescription conditions for veterinary drugs, including antibiotics. The circular also included a roadmap of implementation, that started in 2020 for big-scale farms, 2022 for medium-scale farms, 2023 for small-scale farms, and will be implemented in 2025 for household farms (MARD 2020).

These regulations also impact chicken production. The latter is a growing sector in Vietnam (Cesaro, Duteurtre, and Huong 2020) due to its ease of production, low investment requirements, and high consumer demand. Three types of chicken production systems are commonly defined: integrated farms, family commercial farms, and household farms, with small-scale farms being the most common (GSOV 2016; Bâtie et al. 2022). Interestingly, the different production systems were found to be strongly associated with the antibiotics and alternative feed additives value chain (Bâtie et al. 2022).

Regulations are one of the most common solutions developed to attempt to reduce antibiotic use. However, policy implementation is influenced by both internal and external factors (Brugha 2000) and needs to be adapted to the local context to be effective. Other studies in Vietnam have previously described the antibiotic value chain and policy development organization (Brunton et al. 2019; Pham-Thanh 2020). But interactions between stakeholders have not yet been explored within the context of new regulation implementation and these interactions can have an impact on implementation and must be studied. A system approach provides a comprehensive understanding and identifies which part of the system is likely to be impacted by the change. Furthermore, each stakeholder has unique motivations for complying with new regulations, which must be investigated to identify levers for activation. Stakeholder mapping and analysis (SMA) is a methodology for understanding the implementation context of a policy to assess the feasibility of its application and enforcement (Zimmermann and Maennling 2007; Poupaud et al. 2021; Bordier et al. 2018).

The purpose of this study is to understand how the regulations governing the progressive ban of antibiotics in feed (decree n°13/2020/ND-CP) and on the mandatory prescriptions (circular n°122020TT-BNNPTNT) are understood, accepted, and applied by the different stakeholders in the antibiotics and alternative feed additives value chain. It also provides recommendations for improving their implementation while minimizing the impact on the most vulnerable members of the community.

## Material and methods

We conducted an iterative mapping analysis of stakeholders (stakeholders are described in Table 1) involved in the antibiotics and alternative feed additives value chain and concerned by regulations to reduce antibiotic use in livestock production in Vietnam. We focused on two new regulations that took effect in 2020: the progressive ban of antibiotics in feed (n°13/2020/ND-CP) and the mandatory prescription to buy drugs (n°122020TT-BNNPTNT). These regulations were chosen based on a preliminary study (Bâtie et al. 2022) due to their recent implementation and potential impact on the reduction of antibiotic usage in Vietnam.

**Table 1:** Socio-demographic characteristics of the respondents of the semi-structured interviews (n=39), 2021, Vietnam.

| Characteristics | Number (n=39) | Characteristics | Number (n=39) |
| --- | --- | --- | --- |
| *Gender* |  | *Category of stakeholders* |  |
| Male | 27 | DAH | 1 |
| Female | 12 | DLP | 2 |
|  |  | SubDAHLP | 5 |
| *Location* |  | DARD | 1 |
| North | 28 | Veterinarian district station | 1 |
| South | 11 | Communal veterinarian | 2 |
| *Sector* |  | National research center | 1 |
| Public sector | 13 | Feed company | 3 |
| Private sector | 22 | Alternative feed additives company | 3 |
| International partner | 4 | Company veterinarian | 1 |
| *Territorial level* |  | Importer/Producer | 3 |
| Supra-national | 8 | Distributor | 2 |
| National | 10 | Agencies level 1 and 2 | 5 |
| Provincial | 10 | Private veterinarian | 1 |
| District/communal | 11 | Integrated farm | 1 |
|  |  | Family commercial farm | 2 |
|  |  | Household farm | 1 |
|  |  | International organization | 1 |
|  |  | Foreign research center | 2 |
|  |  | Foreign development agencies | 1 |

MARD: Ministry of Agriculture and Rural Development; DAH: Department of Animal Health; DLP: Department of Livestock Production; Sub-DAHLP: Sub-Department of Animal Health and Livestock Production; DARD: Department of Agriculture and Rural Development, agencies level 1 and 2: local drugstores.

SMA is a method to assess how future policies will be understood by the impacted stakeholders or to support their implementation by identifying key stakeholders, characterizing them, describing their interactions, and exploring their technical and social capital regarding the future policy and their level of influence on its implementation. In detail, it provides relevant information to support the formulation of recommendations to improve the effectiveness of these future policies (Zimmermann and Maennling 2007; Godakandage et al. 2017; Mayers 2005; Reed et al. 2009).

The SMA was conducted in three steps (Bordier et al. 2018; Zimmermann and Maennling 2007; Poupaud et al. 2021): (i) mapping of the antibiotics and alternative feed additives value chain to determine the structural position of the stakeholders within the chain; (ii) analyzing the technical and social capital of the identified stakeholders regarding the regulations to reduce antibiotic use; and (i) identifying the factors influencing the implementation of the new regulations.

### Data collection

Data collection was conducted by four researchers from the Vietnam National University of Agriculture (VNUA) having either a background in sociology or in veterinary medicine, and one researcher from Cirad with a background in veterinary medicine. All the researchers were experienced in participatory epidemiology and/or in conducting qualitative studies and were familiar with the Vietnamese veterinary and animal production context.

We first organized a focus group discussion (FGD) in December 2020 in Hanoi with 12 stakeholders from the antibiotics and alternative feed additives value chain belonging to different categories. Four participants were from the public sector (three from the Sub-Department of Animal Health and Livestock Production (Sub-DAHLP) and one from the veterinarian district station), while eight were from the private sector. The different categories of stakeholders were identified through semi-structured interviews in a preliminary study (Bâtie et al. 2022). The participants were invited based on the list of contacts of one researcher of the team. The objectives were to collectively map the antibiotics and alternative feed additives value chain and identify key stakeholders. The discussion was organized around three topics: (i) identifying the stakeholders in the chain and their roles, (ii) drawing the chain and their interactions, and (iii) identifying existing antibiotic-related legislation [Supplementary Material 1]. The four researchers from VNUA led the three-hour in-person focus group in Vietnamese, using participatory epidemiology tools such as flowchart building (Catley, Alders, and Wood 2012). The first author was present remotely and received simultaneous Vietnamese English translation from a team member. The FGD was recorded, the minutes were transcribed and translated into English, and the value chain drawn by the participant was translated and copied into a Word document.

From March to October 2021, we conducted semi-structured interviews in northern and southern Vietnam with representatives of the categories of stakeholders identified during the FGD. From March to October 2021, 39 interviews were conducted with participants from four provinces (Hanoi, Hung Yen, Hai Phuong, and Bac Ninh) in north Vietnam and three provinces (Hô Chi Minh, Long An, and Dong Thap) in south Vietnam. Participants included government officials (n = 12), private sector workers (n = 18), and foreign partner institutions (n = 4). Table 1 describes the participants. Participants were invited based on the list of contacts of the research team and through an iterative process. Indeed, new categories of stakeholders were added as we deepen our analysis and that new elements have emerged from the semi-structured interviews (Reed et al. 2009). The interview guide was divided into four main topics: (i) position and role within the antibiotics and alternative feed additives value chain, (ii) interactions with other stakeholders and the nature of those interactions, (iii) knowledge and opinions on antibiotic resistance in Vietnam and about the NAP, and (iv) barriers and motivations for implementing the two new regulations under consideration and their expected impacts on the chain [Supplementary Material 2]. Because of the COVID-19 health crisis, five interviews were conducted in person, some were conducted half in person and half online and other fully online. During the interview, a map of the veterinary drug value chain was shared with the participant. The first version of the map presented was built from the results of the FGD and then modified as the interviews progressed. Participants were asked to identify themselves on the map and to describe the nature of their interactions with the other stakeholders. They were able to propose changes to the map. The interviews were audio-recorded and lasted between 60 to 90 minutes. The first author conducted them in English or French, with the research assistant translating into Vietnamese when necessary. Throughout the interviews, notes were taken. Depending on the interviews, field notes were transcribed and translated in English and were expanded with audio recording of the discussion in Vietnamese and completed with the French or English translation, or transcribed and translated in English through verbatim transcription. Before starting the FGD and interviews, the objectives of the study were explained to the participants, and they gave their informed consent to participate in the study. Ethical approval was granted from the Ethics Review Board for biomedical research at Hanoi University of Public Health with the application number 020-419/DD-YTCC.

### Data analysis

The maps created during the FGD, and semi-structured interviews were compiled and reproduced using a flowchart maker and online diagram software (diagrams.net). The semi-structured interviews were analyzed and coded by thematic analysis following the steps described by Castleberry and Nolen (Castleberry and Nolen 2018). We used the qualitative software package NVivo to facilitate the management and organization of our data (V.12,2, QSR International, USA). Thematic analysis is an inductive approach to identifying and analyzing patterns within the transcripts. Codes are descriptive labels attached to similar words or sentences in raw data. They are then reassembled into themes, allowing us to get a bigger picture of the data regarding the research question. Themes were defined *a priori* based on the guide of the interviews. Themes were refined and new ones emerged through a process involving both forward and backward methods. The first author conducted the coding, and the team validated the codebook.

The thematic analysis allowed us to first confirm and improve the map of the antibiotics and alternative feed additives value chain, identify the structural position of the stakeholders, and define their role within this chain. Secondly, each stakeholder who was interviewed was analyzed based on three themes related to their technical and social capital in relation to regulation: legitimacy (institutional position, involvement in the law design, formal or informal flow of antibiotics), resources (knowledge and technical capacities to implement the regulations), and connections (number and quality of relationship) to other stakeholders in the chain (Figure 1) (Zimmermann and Maennling 2007; Poupaud et al. 2021). We were able to categorize each actor according to their capital level by assigning a score ranging from absent (-) to strong (+++). Each category of stakeholders was assigned the average score of participants within that category. Third, we identified the factors influencing the implementation of the new regulations using the chain map, stakeholder classification, and semi-structured interview coding (barriers, and levers to implement the new regulations). Data were triangulated (Olivier de Sardan 2008) through multiple sources of information (different respondents, flow chart), observation when possible, and team discussion.

**Figure 1:**
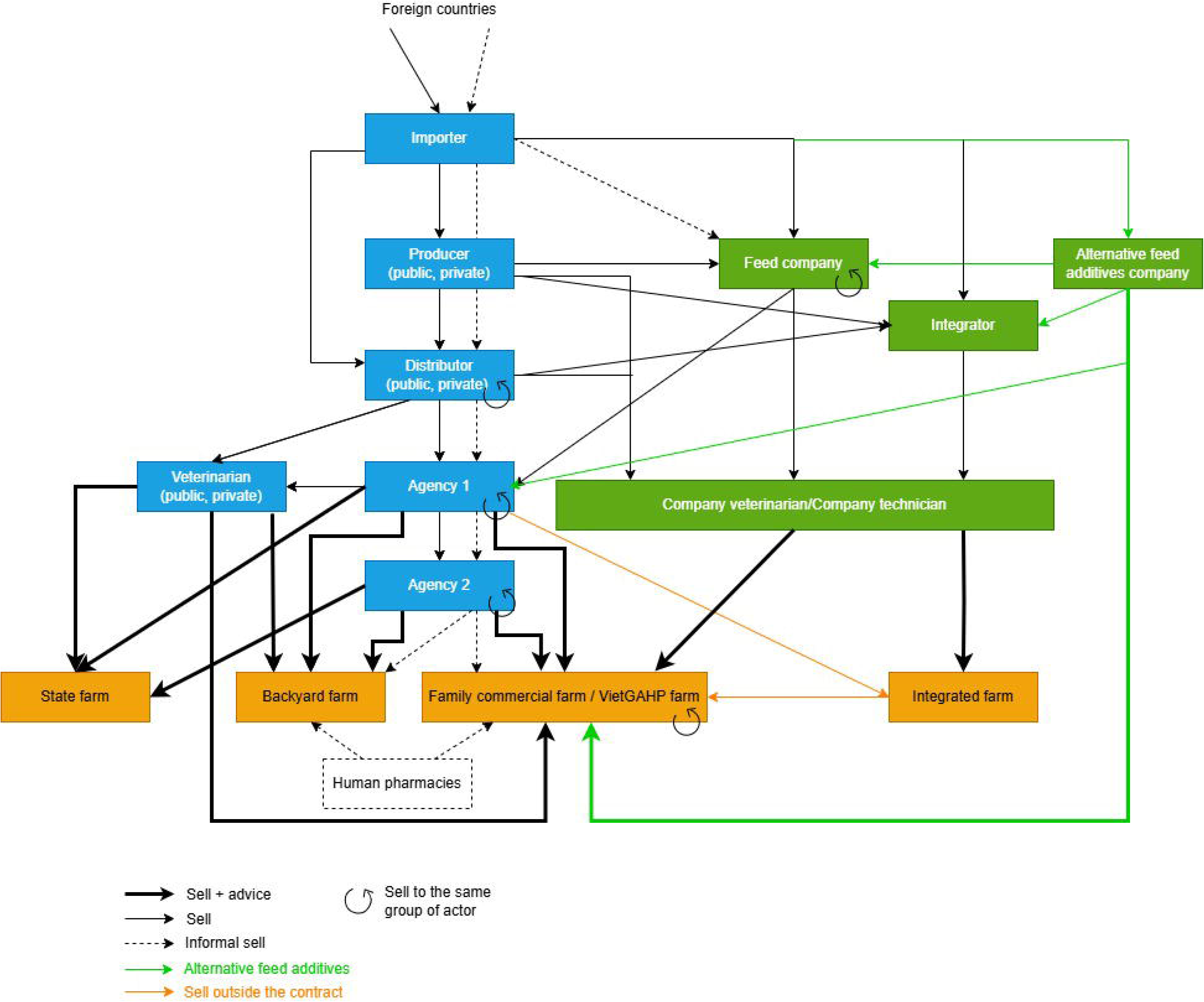
Organization of the antibiotics and alternative feed additives value chain from focus group discussion (n=12) and semi-structured interviews (n=39), 2020-2021, Vietnam. VietGAHP: Vietnamese Good Animal Husbandry Practices; agencies level 1 and 2: local drugstores.

## Results

The results are presented in three parts: 1) structural position of the stakeholders; 2) technical and social capital of stakeholders; and 3) factors influencing the implementation of new regulations.

### 1) Structural position of the stakeholders within the veterinary drug value chain

The veterinary drug value chain, as mapped during the FGD, was revised three times during semi-structured interviews. Figure 1 shows the result of the mapping. From our interviews, there was no difference in chain structure between respondents from North and South Vietnam. Finished products are imported or made in Vietnam using imported components. They are then distributed either directly to farmers or through agencies. An agency was defined by the participants as a shop selling drugs, agricultural equipment, or animal feed and owned by a veterinarian. Agencies 1 and 2 differ in size, source of supply (level-1 agencies are directly connected to drug companies, veterinarians and farmers and sometimes provide level-2 agencies, whereas level-2 agencies target mostly small-scale farmers). Both types of agencies must be run by a veterinary technician that owns a veterinary college or an intermediate degree representing two to three years of study or by a veterinarian with a university degree. They often receive the help of their spouse or other personnel that does not necessarily have the qualifications. In our study two agencies level-1 were run by veterinarians that have a doctorate in veterinary medicine and two were run by veterinary technicians. A third level was occasionally reported by participants, but as it differed between interviews, it was not included in the figure. Private veterinarians in Vietnam are stakeholders called veterinarians by other participants or themselves, with a university, college degree, or without any degree but with some experiences in veterinary medicine. The end users were identified as three types of farms with different input and output sources: integrated farms (contracts with integrators), family commercial farms (private farms), and household farms (backyard farms). VietGAHP (Vietnamese Good Animal Husbandry Practices) farms were certified under the same-named standards. We defined veterinarians as veterinarians who worked for an agency or independently that provided farmers with direct advice and drugs. Company veterinarians or technicians were stakeholders employed by a company (pharmaceutical, feed, alternative feed, or integrator) with a university degree in veterinary medicine or college degree in a technique (veterinary technician, feed technician, …). Their roles are described in Supplementary Material 3.

Interactions among the identified stakeholders varied. The agencies charge a fee for drugs, feed, and materials, but they also offer advice to farmers. These interactions can be facilitated through a contract between farmers, agencies, and distributors. In this case, these agencies receive preferential pricing and benefits if they purchase or sell a certain number of drugs. The interactions can also take place across a network of farms, in which a technician employed by an integrator provides the farm with drugs and advice.

This chain is regulated by authorities at various territorial levels, as shown in Figure 2. The Department of Animal Health (DAH) regulates antibiotic importation, production, and distribution, as well as medicated feed mills. The department is also responsible for delivering practice certificates to veterinarians. The Department of Livestock Production (DLP) regulates feed companies. The DAH oversees the provincial Sub-Department of Animal Health (Sub-DAH), which merged with the Sub-Department of Livestock Production (Sub-DLP) to form the Sub-Department of Animal Health and Livestock Production (Sub-DAHLP). The DAH provide Sub-DAHLP with information and guidelines for implementing legislation. In exchange, they receive reports on the situation in each province. The two last administrative levels are veterinary district stations and communal veterinarians, which operate on the district and communal levels, respectively. In Vietnam, communal veterinarians are stakeholders mandated by the government and are responsible for the sanitary situation, disease control and vaccination campaign. Of the two communal veterinarians interviewed, one did not hold a university or college degree in veterinary medicine. They both had a side-job: private veterinarian or working in another field. Finally, international organizations, international collaborators, and national and foreign research centers work together with government authorities to provide evidence for the development of new policies and technical guidance.

**Figure 2:**
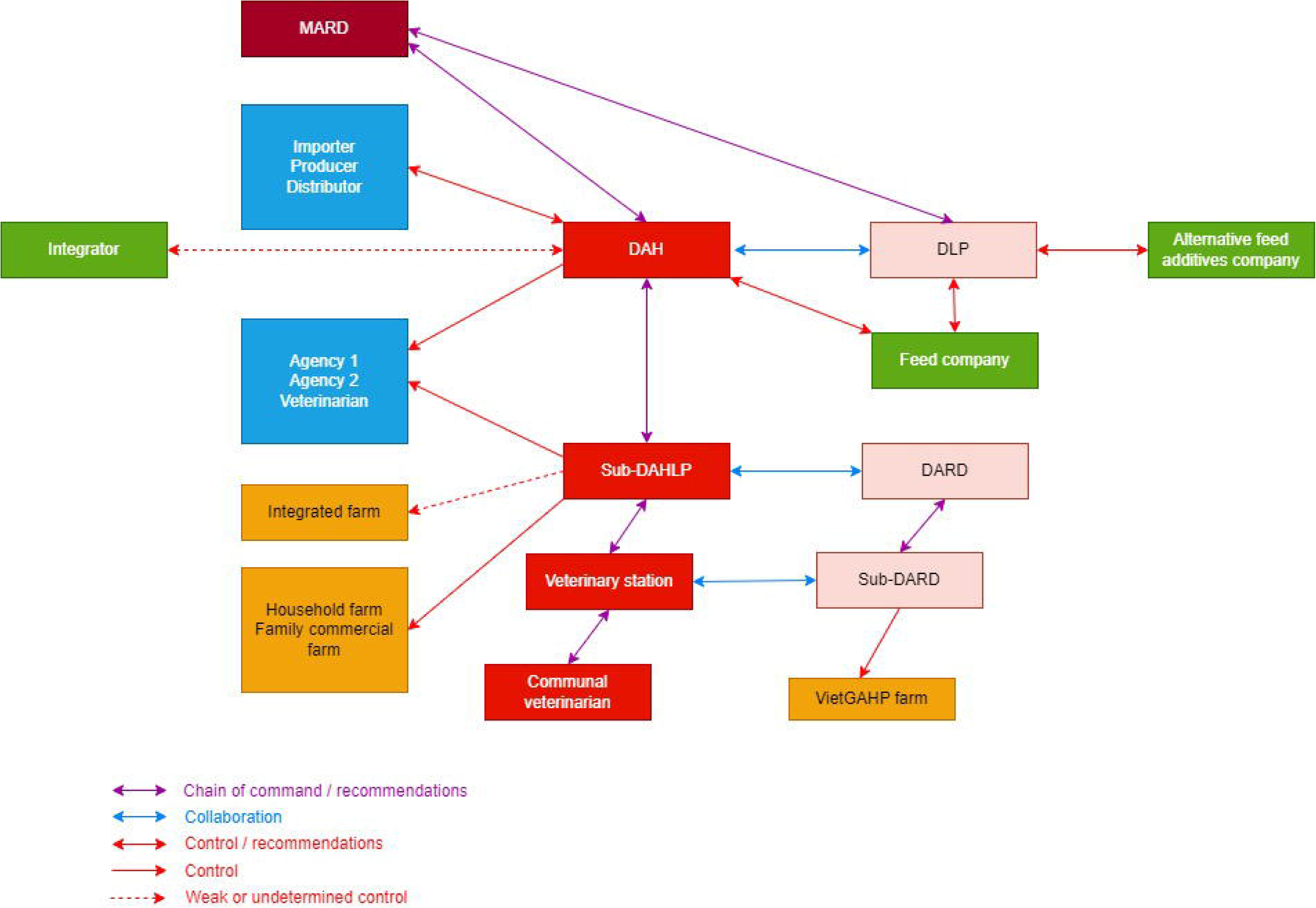
Chain of control of the antibiotics and alternative feed additives value chain and chain of command between governmental authorities from focus group discussion (n=12) and semi-structured interviews (n=39), 2021, Vietnam. MARD: Ministry of Agriculture and Rural Development; DAH: Department of Animal Health; DLP: Department of Livestock Production; Sub-DAHLP: Sub-department of Animal Health and Livestock Production; DARD: Department of Agriculture and Rural Development; agencies level 1 and 2: local drugstores; VietGAHP: Vietnamese Good Animal Husbandry Practices.

### 2) Technical and social capital in stakeholders regarding regulations

Through the FGD and individual interviews, we identified 30 categories of stakeholders affected by the regulations. They were divided into three groups: the public sector comprising governmental bodies and national research centers (n=9), the private sector involving stakeholders from importers to final antibiotic users (n=15), and international partners (n=6) (Figure 3). Analyzing the main functions of the potential key stakeholders helped us identify those who have significant influence. The stakeholders’ technical and social capital based on their legitimacy, resources, and connections is outlined in Table 2 and visualized in Figure 4. More information can be found in Supplementary Material 3. Technical and social capital are two concepts used in sociology as usable resources, skills, abilities and power by individuals (Brock, Kvasny, and Hales 2010). In particular, social capital refers to the set of resources that an individual can mobilize related to he/she permanent more or less institutionalized relationship network (Bourdieu 1980).

**Figure 3:**
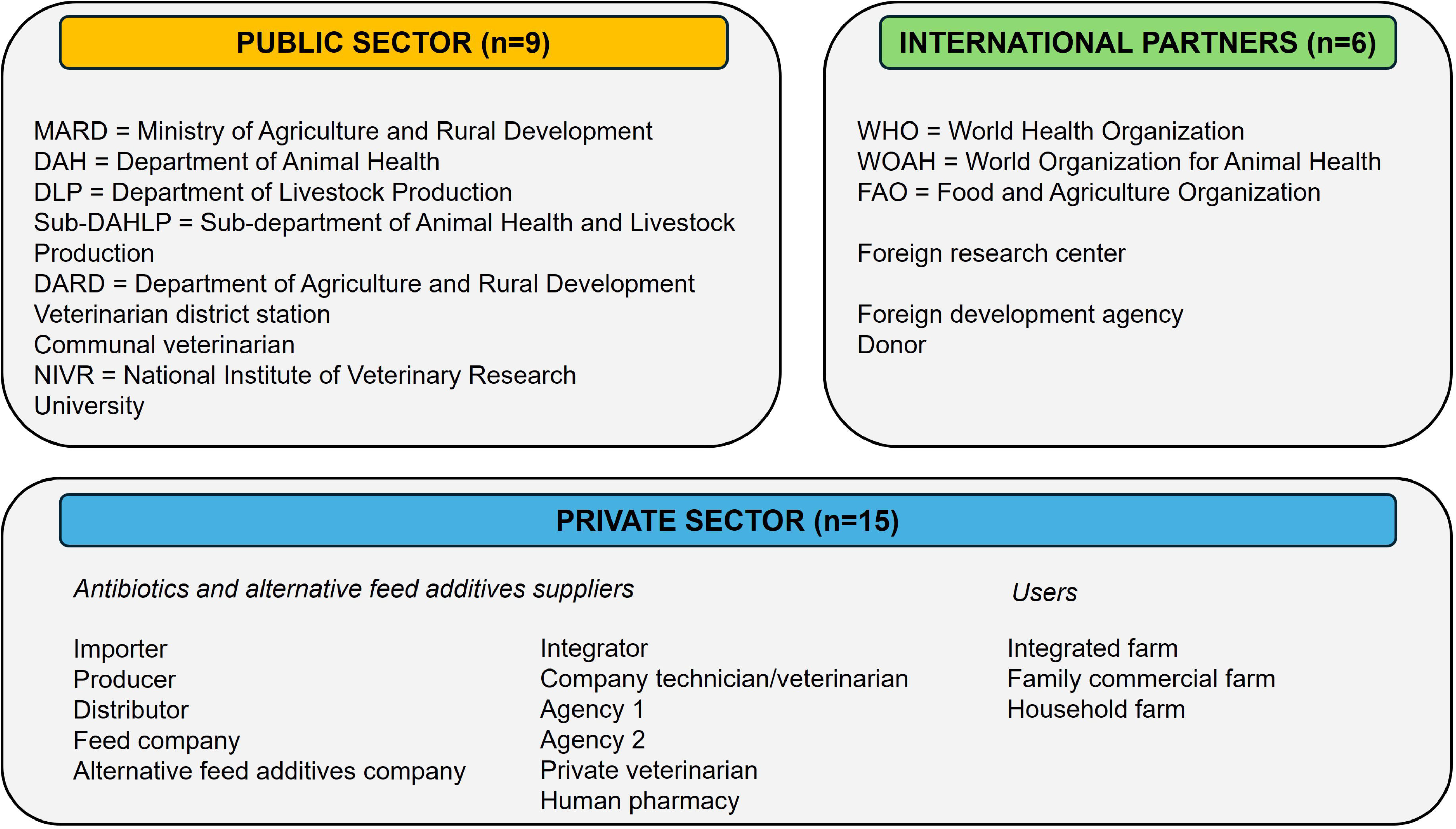
Categories of key stakeholders (n=30) involved in the antibiotics and alternative feed additives value chain from focus group discussion (n=12) and semi-structured interviews (n=39), 2020-2021, Vietnam. Agencies level 1 and 2: local drugstores; private veterinarians: stakeholder called veterinarian by other participants or himself, with a university, college degree, or experiences in veterinary medicine; company technician: stakeholders holding a technical degree in the relevant field of expertise and working for a company; company veterinarian: veterinarian employed by a company; communal veterinarian: stakeholders mandated by the government to report diseases outbreak and manage vaccination programs.

**Figure 4:**
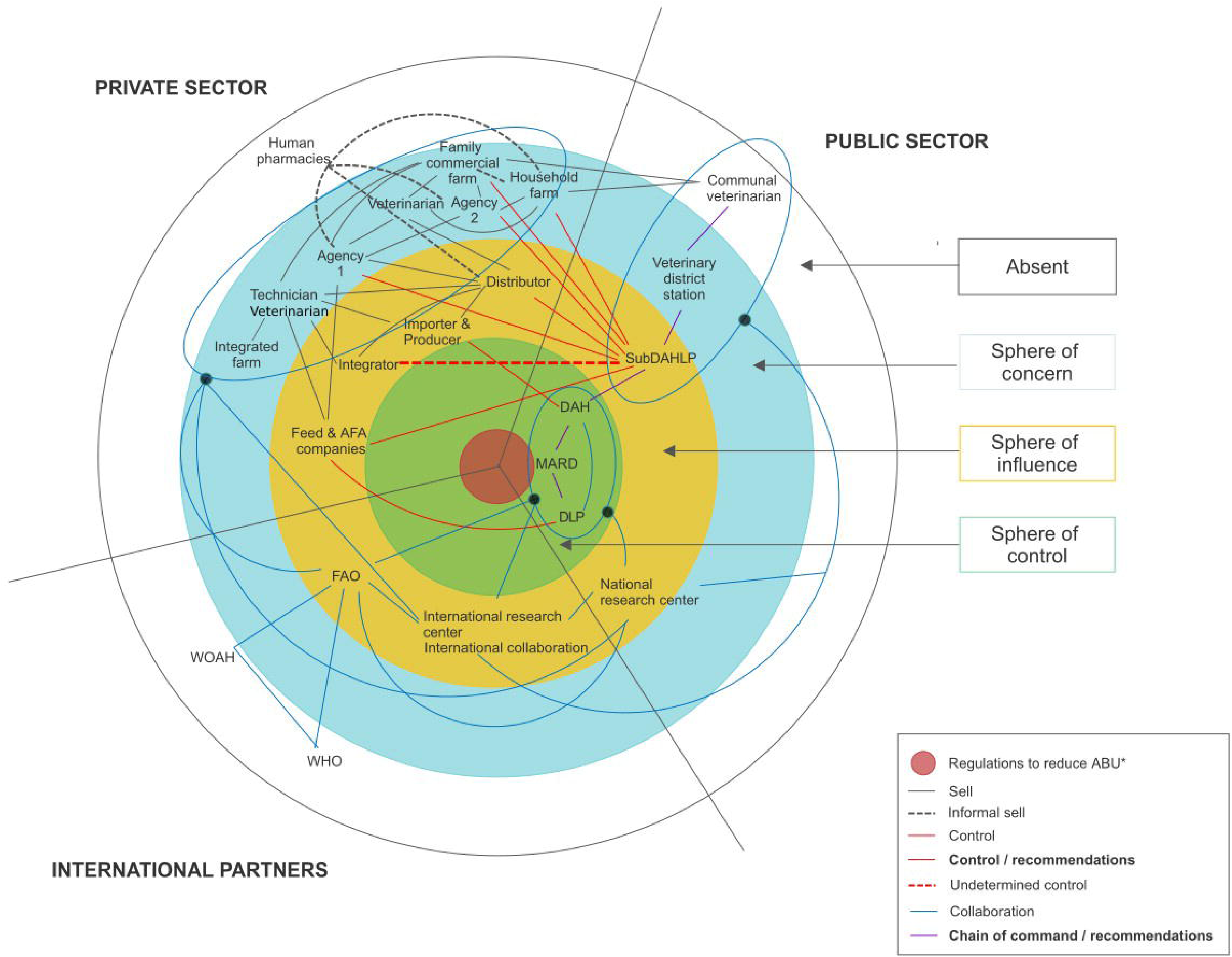
Onion ring visualization of the technical and social capital of the identified stakeholders within the chain regarding regulations to reduce antibiotic use from semi-structured interviews (n=39), 2021, Vietnam. WOAH: World Organization of Animal Health; WHO: World Health Organization; FAO: Food and Agriculture Organization; MARD: Ministry of Agriculture and Rural Development; DAH: Department of Animal Health; DLP: Department of Livestock Production; Sub-DAHLP: Sub-Department of Animal Health and Livestock Production; agencies level 1 and 2: local drugstores.*: focus on new regulations on the progressive ban of antibiotics in the feed (n°13/2020/ND-CP) and mandatory prescription (n°122020TT-BNNPTNT). Sphere of control: stakeholders who are central to behaviors and activities. Sphere of influence: stakeholders who have the power to change the outcome on impacts. Sphere of concern: stakeholders who have no control over the actions of others but who are affected by the regulations. Absent: stakeholders who have no control over regulations. Onion ring visualization adapted from (Bordier et al. 2018; Zimmermann and Maennling 2007).

**Table 2:**
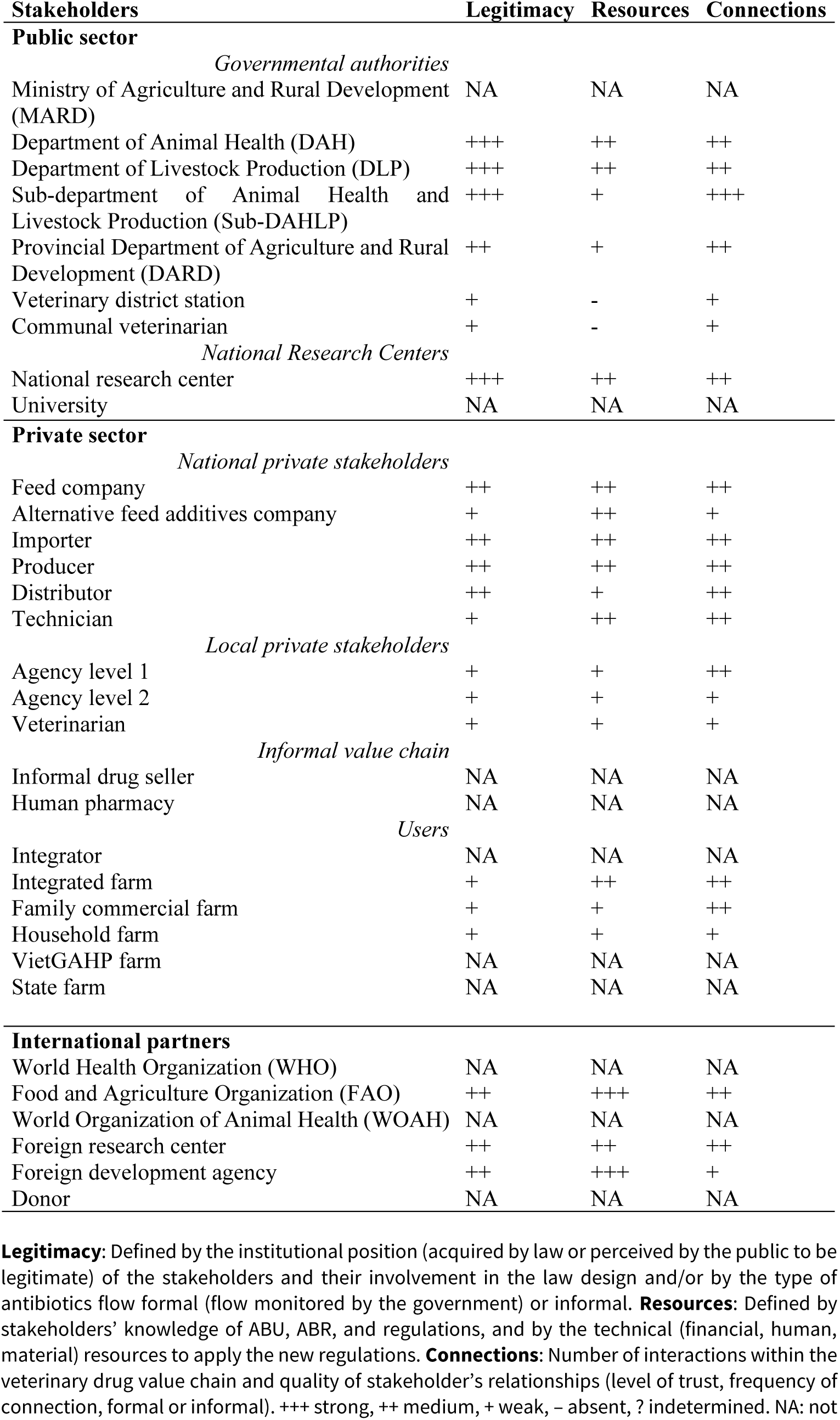
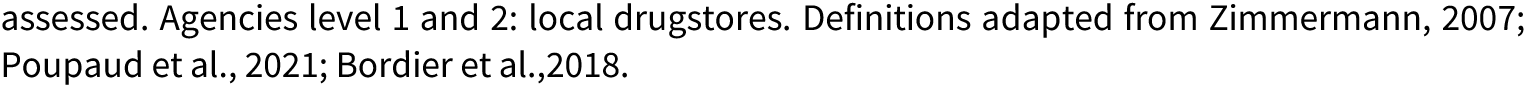
Classification of the stakeholders of the veterinary drug value chain interviewed according to their level of legitimacy, resources, connections regarding new regulations on the progressive ban of antibiotics in the feed (n°13/2020/ND-CP) and mandatory prescription (n°122020TT-BNNPTNT), 2021, Vietnam.

| Stakeholders | Legitimacy | Resources | Connections |
| --- | --- | --- | --- |
| **Public sector** |  |  |  |
| *Governmental authorities* |  |  |  |
| Ministry of Agriculture and Rural Development (MARD) | NA | NA | NA |
| Department of Animal Health (DAH) | +++ | ++ | ++ |
| Department of Livestock Production (DLP) | +++ | ++ | ++ |
| Sub-department of Animal Health and Livestock Production (Sub-DAHLP) | +++ | + | +++ |
| Provincial Department of Agriculture and Rural Development (DARD) | ++ | + | ++ |
| Veterinary district station | + | - | + |
| Communal veterinarian | + | - | + |
| *National Research Centers* |  |  |  |
| National research center | +++ | ++ | ++ |
| University | NA | NA | NA |
| **Private sector** |  |  |  |
| *National private stakeholders* |  |  |  |
| Feed company | ++ | ++ | ++ |
| Alternative feed additives company | + | ++ | + |
| Importer | ++ | ++ | ++ |
| Producer | ++ | ++ | ++ |
| Distributor | ++ | + | ++ |
| Technician | + | ++ | ++ |
| *Local private stakeholders* |  |  |  |
| Agency level 1 | + | + | ++ |
| Agency level 2 | + | + | + |
| Veterinarian | + | + | + |
| *Informal value chain* |  |  |  |
| Informal drug seller | NA | NA | NA |
| Human pharmacy | NA | NA | NA |
| *Users* |  |  |  |
| Integrator | NA | NA | NA |
| Integrated farm | + | ++ | ++ |
| Family commercial farm | + | + | ++ |
| Household farm | + | + | + |
| VietGAHP farm | NA | NA | NA |
| State farm | NA | NA | NA |
| **International partners** |  |  |  |
| World Health Organization (WHO) | NA | NA | NA |
| Food and Agriculture Organization (FAO) | ++ | +++ | ++ |
| World Organization of Animal Health (WOAH) | NA | NA | NA |
| Foreign research center | ++ | ++ | ++ |
| Foreign development agency | ++ | +++ | + |
| Donor | NA | NA | NA |

**Legitimacy:** Defined by the institutional position (acquired by law or perceived by the public to be legitimate) of the stakeholders and their involvement in the law design and/or by the type of antibiotics flow formal (flow monitored by the government) or informal. **Resources:** Defined by stakeholders' knowledge of ABU, ABR, and regulations, and by the technical (financial, human, material) resources to apply the new regulations. **Connections:** Number of interactions within the veterinary drug value chain and quality of stakeholder's relationships (level of trust, frequency of connection, formal or informal). +++ strong, ++ medium, + weak, - absent, ? indetermined. NA: not
assessed. Agencies level 1 and 2: local drugstores. Definitions adapted from Zimmermann, 2007; Poupaud et al., 2021; Bordier et al., 2018.

#### Public sector

The public sector included government authorities and national research centers.

##### Government authorities

Their legitimacy, resources, and connections decreased throughout the value chain. Respondents indicated that the central level was critical in the development of the NAP and regulations (Figure 4). The DAH was described as an “umbrella” that manages all activities related to the veterinary drug value chain, and in the NAP, they are identified as the focal point for implementation. The central government (DAH and DLP) had a good understanding of ABR, ABU, and regulations, though they acknowledged that they were not always aware of the full ABR situation in Vietnam.

> “I don’t know exactly what the current situation is, many projects are working on antimicrobial resistance. (…) actually, I don’t believe we have a full picture for the country.” (Interview 12 (IW12), government authority)

The DAH and DLP also collaborated to develop the circular on mandatory prescriptions (circular 12). They are linked to a number of stakeholders (MARD, international partners, Sub-DAHLP, national private stakeholders), but not directly to small holders and drug sellers locally. The central government’s legitimacy, resources, and connections provided it influence over the legislative process and the formulation of new regulations.

The Sub-DAHLP was viewed as a link between central, district, and communal authorities. Their missions were to monitor everything related to antibiotic use, including drug agency control (certificate of practice, use of banned substances, and quality testing). However, participants suggested that their mission had limitations as they are too few to control all the shops.

The Sub-DAHLP could also offer suggestions for the development of new regulations. Other stakeholders rated their knowledge as average, and in some provinces, they were perceived to lack the human and technical capacity needed to enforce new regulations. Indeed, differences between provinces were observed in the five interviews conducted with Sub-DAHLP representatives, not only in terms of ABR and regulation knowledge, but also in technical capacity to enforce the regulations. The Sub-DAHLP was described as having a close relationship with all stakeholders in the veterinary drug value chain, particularly with drug agencies. Sub-DAHLP employees are responsible for controlling agencies, but they also frequently own them as a secondary source of income. A respondent raised the possibility of a conflict of interest between the competent authority and the private sector in terms of ABU management. The Sub-DAHLP also plays an important role in ABU and ABR research. They not only authorize data collection, but they also connect research projects, farms, and drugstores.

> “What also surprised me is the link between the Sub-DAH vet and the drugshops. A lot of them, at least 30/50%, are owned by staff affiliated with Sub-DAH or former Sub-DAH staff.” (IW28, international partner)

District veterinarians at the veterinary district station were not perceived to play a significant role in Vietnam’s antibiotic reduction regulations. Other stakeholders perceived them to have limited knowledge of ABU and ABR, as well as limited capacity to implement regulations. They were also described as relaying information about new regulations to farmers and drug sellers.

Communal veterinarians were perceived to have no role in regulatory development or implementation; their primary function was to organize mandatory vaccination. One of two communal veterinarian interviewees was not qualified veterinarians and had limited knowledge, learning from the experiences of drugstore agents and farmers. They admitted to not being very involved in the fight against ABR. Their primary source of income was treating diseases in farmers’ livestock and selling antibiotics; the second came from government subsidies. They are directly connected with farmers in the local area.

##### National research centers

National research centers were considered as knowledgeable about the ABU and ABR situation in Vietnam. A researcher working in a foreign research center perceived them as getting more involved in the fight against ABR. By leading research on ABU, ABR, and antibiotic residues, they were providing evidence that authorities used to create regulations. They were directly overseen by the MARD, either DAH or DLP. They also had strong ties to international organizations and foreign research institutions. They said that they had frequent contact with farmers and drug sellers while conducting research. They were not, however, directly connected to MARD; rather, their findings were reported to MARD by international organizations.

#### Private sector

The private sector included stakeholders operating on a national and local scale, as well as users.

##### Private national stakeholders

This category included drug, feed, and alternative feed additive companies and integrators.

They were perceived by authorities and other private stakeholders as having the necessary knowledge, technical capabilities, and human resources to implement the regulations. In our study, companies were operating at provincial, national or supra-national levels. These companies employed veterinarians and veterinary technicians who provide free technical advice to large-scale farmers on a contract basis. They have studied veterinary medicine at the university or college level, so they are authorized to write prescriptions, giving them an advantage in enforcing the regulations. The most influential were involved in the regulation design, while others were aware of regulatory changes before local stakeholders. Before the regulations were issued, some drug companies began producing alternative medicines, and feed companies replaced antibiotics with alternative supplements. Alternative feed additive companies provided large-scale farms with free guidance as a trial. However, they reported insufficient resources to deal directly with smallholders, and their products’ packaging was not always appropriate for them.

They had a strong connection that was formalized through a contract with integrated farms, large family commercial farms, and level 1 agencies. They had no direct contact with households or small and medium-sized family commercial farms.

##### Private local stakeholders

This category included level 1 and 2 agencies and veterinarians who did not have a shop. They were perceived as having weak legitimacy because they were not part of the decision-making process when designing regulations and because they sometimes sell banned antibiotics.

> “Most of the vet drug shops in Vietnam sell banned drugs. That’s the reality.” (IW22, agency level 1)

They were also seen as lacking the resources to implement new regulations. Level 1 agency representatives usually had a veterinary degree and an in-depth knowledge of ABU and ABR, but not all level 2 agencies were run by veterinarians, and not all employees were trained in animal diseases and antibiotic use. Other stakeholders described them as being underinformed, and during the interviews, they were not always aware of the new regulations. They were, however, perceived to play an important role in the implementation of regulations due to their close connection with farmers.

> “In some remote areas, people do not have advanced training; this includes veterinarians who do not have practice certificates as they have very little work, with only a few small-scale livestock farms.” (IW23, local veterinary services)

##### Users

The users included integrated farmers, family commercial farmers, and household farmers producing chicken. Users are defined as having a low level of legitimacy because they were not consulted in the law-making process (with the exception of the most important integrated farms), despite being perceived as directly affected by regulatory changes.

According to government authorities, private national and local stakeholders, and researchers, farmers lacked knowledge of regulations and prudent ABU practices. A foreign researcher had, however, nuanced his statement as there were several types of farmers. Small-scale farms were frequently cited as the source of antibiotic misuse in Vietnam.

They had strong connections among them, sharing their experiences. The strength of their connection with agencies differed between users. Some of them went to various agencies without seeking advice, purchasing drugs solely based on their own experience. Others, however, relied heavily on advice from trusted agencies. Integrated farms and large family commercial farms were connected to drug and feed agencies or integrators by a contract.

#### International partners

International partners included international organizations, foreign research centers, and foreign development agencies. International partners provided evidence of the ABU and ABR situation in Vietnam and were thus considered legitimate. They self-assessed their influence as moderate, noting that they were not the final decision-makers and could only make recommendations to policymakers. However, the DAH relies on research project findings to develop new regulations, and the majority of them were involved in the development of the NAP alongside national authorities.

> “I think we have an influence because I have seen a lot of my papers cited even in the legislation (…) However, you don’t know how much influence there is” (IW28, international partner)

International partners were perceived to have a thorough understanding of ABU and ABR in Vietnam. They had the ability to organize workshops, conduct research, create guidelines, and assist the government. Respondents also identified donors as having greater influence because they bring fundings and thus can set the direction of the governmental action.

International partners were described as being indirectly linked to the value chain. Collaboration between different research centers, foreign development agencies, and international organizations was perceived to be multifaceted. Their connections with companies or integrators appeared limited, as conducting research with integrated farms could be difficult. They did, however, collaborate with other companies to conduct product trials.

### 3) Factors influencing the implementation of new regulations

Based on our analysis of the stakeholders’ capital regarding regulations, we identified 10 factors that influenced, positively or negatively or both, the implementation of new regulations [Supplementary Material 4].

#### Stakeholders’ perceptions of the new regulations

The vast majority of value chain stakeholders were in favor of the new regulations but they were not confident in their short-term implementation. They recognized their importance in reducing antibiotic use, fighting antibiotic resistance and protecting human health. Respondents thought that farmers will be forced to change their practices over time, and that new regulations are an important step toward that goal. Except for government stakeholders, all categories of respondents stated that they would be difficult to implement and lacked confidence in their short-term implementation. Respondents also compared the situation in the human sector, where prescriptions are required to purchase drugs, but this regulation isn’t enforced.

> “Without a doctor’s prescription, you can still buy medicine for self-treatment.” (IW39, household farm)

> “This law is a feasible way to reduce antimicrobial use but it will take 5-10 years for farmers to adapt to it. The government expects it to be achieved in 1-2 years, but that’s impossible, it will take a long time to raise farmers’ awareness.” (IW24, local veterinary service)

#### Level of knowledge

Gaps of knowledge about ABU, ABR, and on regulations were perceived as barriers to their implementation.

For the respondents, the rise of ABR was due to overuse of antibiotics, incorrect dosage and duration, and the use of banned antibiotics by farmers. Stakeholders, including farmers reported that the emergence of resistance on farms has led them to increase dosage and switch of antibiotics, leading to untreatable conditions, higher farm expenditure, and the need to find alternative solutions. Only a few respondents identified the impact on human health, and most farmers perceived ABR as invisible with only long-term effects. Except for farmers, all other stakeholder categories perceived users, national and local private stakeholders and local governmental authorities as not having enough knowledge on ABU and ABR.

> “It’s true that the officers don’t really understand what antimicrobial resistance is, not to mention farmers. Especially, they believe what they see, but the effect of AMR has long-term consequence that are not immediately visible.” (IW25, local veterinary service)

In terms of regulations, stakeholders, except for central government actors, had a limited understanding of the decree on antibiotic usage in feed. Their awareness of the circular on prescription was limited and confused with a previous law (Law on Veterinary Medicine, 2015). Farmers and local drug sellers were the least aware of regulations, followed by communal and district veterinarians and sometimes Sub-DAHLP. National companies, the national government, and researchers were all aware of the regulatory changes. Respondents’ understanding of mandatory prescription regulations was low, especially for farmers who did not understand the benefit of this regulation. Mass media, the internet, and community loudspeakers were the principal sources of information for drug sellers and farmers, but they recognized the need to increase communication around new laws. Some workshops were also organized by companies together with the government.

#### Technical guidance availability on the implementation of regulations

At the central and international levels (private, public, and international partners), the respondents were all aware of the regulations but unsure of how to implement them in practice. No technical guidance was provided, and it was not clear how regulations should be enforced. Moreover, regulations were also perceived as incomplete, especially because details regarding on-farm inspections were not included (number of inspections per year, exact procedure). They consider this to be significant because, even if buying feed containing antibiotics for chickens older than 21 days was banned, farmers continued to add antibiotics to the feed. For some respondents, stricter sanctions should also be included in the regulations to oblige farmers and feed producers to comply with them.

The lack of technical guidance and economic interest for farmers was also identified as a barrier to their implementation.

> “If the law is not thorough, difficult to implement, and does not bring economic benefits, it is not mandatory for me to follow it.” (IW36, integrated farmer)

#### Conflict of economic interest between stakeholders

Those who didn’t comply with the regulations were perceived to have an economic advantage over the others, which encouraged non-compliance with regulations. For farmers, using antibiotics for prevention was a way to reduce disease incidence and ensure good production. Buying antibiotics over the counter was a way to save time in case of flock disease, and not respecting the withdrawal time was also done for economic purpose. For drug sellers, selling antibiotics without a prescription is faster than with one, allowing them to sell more drugs and thus increase their income.

> “This law cannot be applied in practice when chickens are sick; I buy medicine myself rather than waiting for a veterinarian prescription.” (IW37, family commercial farmer)

Feed companies that add antibiotics to feed benefit more than others. Indeed, if feed is perceived as less efficient because of the absence of antibiotics, farmers will switch to a feed supplier known to add antibiotics to feed to maximize their production. Respondents reinforce the fact that, for regulations to be effective, they must be applied by everyone at the same time.

> “Enforcement is one of the issues because of the violation of certain (…). If one of them uses it illegally, that means that somehow, they have the benefit of market competitiveness.” (IW34, international partner)

#### Scale-related management discrepancies

The predominance of small-scale farms in Vietnam was perceived by respondents as a major barrier. Stakeholders reported that they had poor farming management practices, which were accentuated by intermittent production. Indeed, some farmers produced a few hundred chickens for the Lunar New Year and then stopped their production after selling the batch. Farmers treated their chickens themselves or according to the experience of other farmers and bought drugs without a prescription based on their own experience. This practice was confirmed by a family commercial farmer that explained not always following the recommendations of the health professional, because they relied more on their own or other farmer’s experience and wanted to treat their animals immediately. However, the household farmer reported calling the veterinarians and the family commercial farmers going or calling the veterinarian for unknown disease. Respondents raised that small-scale farmers aged between 50 and 60 showed less willingness to change compared to the younger generation.

Moreover, there was insufficient control over small-scale farms because the government did not have the human and financial capacity to inspect them all.

> “Even if we don’t put antibiotics in feed, they can still buy drugs from drug agencies and add it themselves.” (IW29, feed company)

Additionally, some drug sellers sold antibiotics without the proper qualifications. This is made possible by the limited control capacity of Sub-DAHLP to inspect all small drugstores.

Integrated and large-scale farms were perceived to be more compliant with legislation. Firstly, they were more aware of regulatory changes and had time to adapt due to their direct link with integrators. Integrators, drug, and feed companies were also better prepared for regulatory change because in-company veterinarians were qualified to write out prescriptions under the new regulation. Also, respondents explained that it was more difficult for a large farm to add antibiotics to feed and to mix them themselves than for small scale farms. Government or contract farm integrators carried out more on-farm inspections due to the products being sold in supermarkets where drug residue tests are performed.

#### Technical and financial barriers to the implementation of regulations at local level

Locally, respondents stressed the technical and financial barriers to implementing the new regulations. First, in some areas, there were not enough certified veterinarians for the number of farmers, especially given the large number of small-scale farms. Thus, due to a shortage of qualified veterinarians, systematic prescriptions for the purchase of antibiotics would be impossible to implement.

> “In some remote areas, people do not have advanced training; this includes veterinarians who do not have practice certificates as they have very little work, with only a few small-scale livestock farms.” (IW23, local veterinary service)

Companies and agencies expressed that small-scale farms lacked adequate biosecurity measures (poor farming management and housing conditions), leading to a high disease incidence and explaining high antibiotic usage in prevention. This was exacerbated by the tropical weather in Vietnam characterized by high temperature, humidity and frequent changes in forecast that increase the infectious pressure. One farmer also mentioned weather change without mentioning biosecurity. A foreign partner also stressed the lack of farming practice expertise of farmers (related to intermittent production) that should be better trained by agencies.

> “The germs are abundant, the density of farming is thick, the climatic conditions are favorable for bacteria to grow, so they meet the pressure of disease.” (IW7, distributor)

Alternative products were perceived as more expensive than antibiotics and available in inappropriate packaging by farmers and other drug sellers. Moreover, small farmers were usually not approached by an alternative company, as stated by one representative of this actor category. Large-scale farms had easier access to alternatives because alternative medicine companies provided free trials and technical advice. However, some farmers and drug sellers reported using garlic, herbs, or vitamins on a regular basis. University research was also underway in one of the studied districts on the use of herbs to improve disease treatment and meat quality.

#### Informal distribution channels

The informal market was perceived as a barrier to the implementation of new regulations. Indeed, unlicensed products from Vietnam were imported across the Chinese border and then sold over to agencies via the internet or distributors. The government lacked capacity to control this illegal flow of antibiotics as informal vendors were perceived to be powerful, while the business generated a lot of money. As a result, some shops sold low-quality (diluted) drugs and banned substances.

> “Counterfeit drugs with less than 70% content are common in Vietnam. This affects vets making prescriptions. Drugs are used according to instructions and are found to be inefficient, so they need to double the amount of drugs, which creates antibiotic resistance.” (IW4, local veterinary service)

Farmers continued to buy banned antibiotics because they were perceived to be more effective. To the contrary, poor-quality drugs were perceived as a major issue that the government should address. Respondents also reported the presence of unlicensed shops run by people without practice certificates.

#### International influence

The Vietnamese government developed its NAP and implemented corresponding regulations, partly to respond to international demand. These regulations were strongly influenced by foreign technical experts and were therefore similar to European regulations.

> “If there is a complete ban, this will be due to trends worldwide and in Vietnam, which is gradually eliminating antibiotics” (IW19, distributor)

However, some respondents had a negative perception of this international influence. Indeed, as Vietnamese breeding conditions differed from those in Europe, they identified that some regulations were poorly suited to the context and difficult to implement.

For companies that must follow the international trend of reducing antibiotics in order to export their products, the international influence was seen as a motivation. Indeed, complying with these regulations was a step toward reaching the food safety standards required for the international market.

#### Consumers demand for food safety

Producing higher-quality food that could be sold for a higher price was a motivation at all levels and in all sectors. Indeed, respondents reported that consumers were more aware of ABR and food quality issues and that farmers had to progressively comply with consumers’ demands. However, because not all consumers had the possibility to pay a higher price for chickens, this lever was still identified as moderate. Household farmers also reported not using antibiotics in their flock to protect their health.

Companies had started to develop labels for chicken produced with less antibiotics or fed with herbs that better suit urban consumer demand. Moreover, the VietGAHP certificate is a standard needed to sell to supermarkets. The requirements to get this certification included, among others, the recording of ABU and the absence of antibiotic residues. Alternative medicine companies were also driven by the desire to create safer products.

> “I know that some companies have really made it part of their strategy to reduce the use of antibiotics. So we’re going to discuss how to support this via our solutions and farm management. (…) Some integrators are aiming for zero antibiotic in line with European practices.” (IW32, alternative feed additives company)

#### Reducing the burden of antibiotic resistance

Reducing the burden of ABR to protect public health was a motivation shared by governmental authorities (up to the provincial level), researchers, and a variety agencies, distributors, and alternative medicine companies. By developing the antimicrobial resistance Sub-Steering Committee, the Vietnamese government has shown that ABR is now considered an important issue. The National Institute of Veterinary Research (NIVR) was identified as playing a central role in this committee. The central government perceived itself as having a responsibility for public health to tackle the ABR threat.

The private sector was also concerned about ABR as a public health issue. But some veterinarians and agency level 1 and 2 representatives explained that reducing the burden of ABR resulted in increased profits for them. Indeed, according to them, farmers using less antibiotics will earn more money and be able to buy more supplies from their shop. Indeed, many agencies sold to farmers on credit. Additionally, mandatory prescriptions were sometimes perceived as an economic advantage for drug sellers. Indeed, farmers will have to rely on veterinarians to use the correct dosage of drugs, which will lead not only to accurate advice to limit the development of resistance but also an increase in their income. Using less antibiotics was also associated with reducing production costs for farmers and integrated companies.

## Discussion

This study aimed to understand whether stakeholders in the veterinary drug value chain in Vietnam understood, accepted, and implemented regulations to reduce ABU, with a focus on the ban of antibiotics in feed (decree n°13/2020/ND-CP) and mandatory prescription (circular n°122020TT-BNNPTNT). To that end, we first obtained a comprehensive representation of the antibiotics and alternative feed additives value chain, as well as a description of the interactions between the different stakeholder categories. We distinguished between stakeholders operating at the national level and the local level. We then identified the technical and social capital of each stakeholder in relation to the regulations designed to reduce ABU. We showed that the level of understanding, acceptance, and implementation of the new regulations varied according to the structural position and influence of the stakeholders within the chain. The level of influence, which depends on their level of legitimacy, resources, connections, and roles in the legislative process, seemed to be higher for drug companies, feed companies, alternative feed additives companies, and integrators than for farmers, level 1 and 2 agencies, and veterinarians. Finally, the first two steps allowed us to identify 10 factors that had a neutral, negative, or positive influence on the implementation of the new regulations. According to the respondents, the high proportion of small-scale farms represented one of the main barriers to their implementation, whereas large-scale production systems seemed to be the most adaptable to regulatory change and thus the least directly impacted. Indeed, small-scale production systems still predominate in Vietnam, but in recent years the number of large farms has increased (Bâtie et al. 2022; GSOV 2016). At the local level, a lack of knowledge about ABU and ABR, the regulations themselves, and their objectives were reported to be important barriers. Finally, enablers to implement regulations were concerned about ABR as a health risk and food safety issue for all stakeholders.

### Advantages and bias of conducting an iterative stakeholder mapping and analysis

In this study, we explored the structural position and technical and social capital of the stakeholders regarding the regulations related to ABR through a systemic (Peters 2014) and interdisciplinary approach to develop recommendations adapted to the Vietnamese context (Kakkar et al. 2018). We aimed to translate policy changes into changes of practice that would effectively reduce the use of antibiotics in chicken production (Baudoin, Hogeveen, and Wauters 2021). To accomplish this, we conducted an iterative stakeholder mapping and analysis (SMA). Our process was iterative as the data collection and data analysis were deepened until all categories of stakeholders have been identified, their positions and relationships mapped and their motivations and barriers explored (Varvasovszky and Brugha 2000). Participatory approaches helped to reduce communication barriers and stimulate discussion on particularly sensitive topics (Bordier et al. 2018). This approach has already been applied to ABR surveillance programs in Vietnam (Bordier et al. 2018) and the antibiotic value chain in Laos (Poupaud et al. 2021). By exploring barriers to the implementation of new regulations using a participatory approach, we identified systemic lock-ins (economical, political, technical, and social) to reducing antibiotic usage in chicken production in Vietnam (Baudoin, Hogeveen, and Wauters 2021).

However, we recognize that our study has some limitations. To begin, almost all of the interviews were conducted online due to the health crisis. This format present a great interest in term of time and budget and no significant difference in thematic content (length of the interviews, number of words, number of codes generated) with in-person interviews have been shown (Krouwel, Jolly, and Greenfield 2019; Namey et al. 2020). However, we have identified several biases that could have hampered our study. Direct observations are reduced, and some information is lost (lack of informal discussions, misunderstandings, longer time to build trust, and more complicated debriefing with the team). Another limitation may have been the sensitivity of the subject to the participants. Indeed, these regulations are already enforced or will be soon, and participants might have been worried about the consequences of expressing their true opinions. We attempted to reduce this bias by interviewing several respondents from the same category and gathering information from the perspective of other participants. However, we failed to contact informal stakeholders who could have helped us further describe the informal value chain. Due to time and technical constraints, we only interviewed one representative for some categories of stakeholders. Thus, it might have been relevant to interview another veterinarian district station to clarify their role in the drug value chain. The identification of stakeholders was also driven by the knowledge and experience of the Vietnamese context of the authors. However, the iterative process allowed us to go beyond this limit as new participants were recruited based on the participant’s knowledge. Finally, translation might also have caused misunderstandings. We are also aware that the representation of the veterinary drug value chain may be incomplete, as it is based on stakeholder interviews and reflects the situation at the time of data collection in 2021. We can however assume that at the time of publication, the results will not be significantly different because as the participants pointed out, human behaviors and policies or regulations are not easy to change in the short term.

The inclusion or exclusion of some categories of stakeholders may influence the conclusions drawn and introduce bias because of their different perspectives on the regulations. Small-scale farmers have been blamed for antibiotic misuse and low regulatory compliance. However, our sample lacked small farmers compared to other stakeholders, and we did not capture their perspectives on their role in ABR as identified by other stakeholders. Thus, additional research is needed to investigate farmers’ perceptions about the ABU reduction and to identify how they adapt to the need for changing practices.

### Influence of stakeholders on the implementation of regulations

The Sub-DAHLP appeared to be the link between the central level and local stakeholders. They had field expertise and were consulted by the upper level, while they transmitted information on ABR, ABU, and regulations to the lower level. They were in contact with drug sellers, veterinarians, and farmers (though they were unable to inspect all farms), as well as researchers. However, respondents reported differences between provinces on regulatory implementation, which might impact the benefits of the regulatory changes. National private stakeholders were consulted on the design of the new legislation, and we saw better understanding, compliance, and adaptability among these stakeholders. They were also better controlled by the competent authorities. They were motivated to reduce their ABU to access the international market and export their products. However, our study was unable to determine the exact influence of private companies on the design of the law. Local stakeholders were not involved in the process and showed a lower level of understanding and compliance. Nevertheless, they were identified as key stakeholders in achieving objectives due to their position in the chain as end-users and must be more involved.

International partners perceived themselves as having a moderate influence on the legislative process. However, in Vietnam, ABU and ABR monitoring remained dependent on research projects, and legislation was partly developed based on the recommendations of international researchers. The NAP was adopted in response to a call from international organizations (OMS 2015) and influenced by international stakeholders, even though the final decision rests with the government. Regulations were sometimes elaborated too far away from Vietnamese reality. Consequently, these regulations developed by national authorities under the influence of international factors may favor production intensification in Vietnam. As explained by one participant, the policy to intensify the farms was done without the farmers’ involvement. To be effective, regulations need to be adapted to the context rather than simply copied from another country’s regulations (Kirchhelle et al. 2020). For example, there were still a large number of small retailers who lacked the legally required practice certificates or qualifications, and they were the only source of antibiotics in remote areas. In this case, strict regulation could limit small-scale farmers’ access to antibiotics.

### Difficulties to implement the regulations

Our study showed that the new regulations would be difficult to implement in Vietnam in the short term, and that local compliance could take longer than anticipated. Indeed, respondents stressed that the use of antibiotics for preventive purposes, by mixing antibiotics themselves with feed, is deeply rooted in the practices of farmers and drug sellers. In addition, these practices are difficult to control due to the large number of small-scale farmers in the country. This finding is consistent with several studies that have shown that the regulatory enforcement of antibiotic usage is low or absent in low- and middle-income countries (LMICs), particularly in Vietnam (Nguyen et al. 2013). The new NAP issued in 2021 also highlighted the need for better regulatory enforcement to fight against ABR (MARD 2021), and an analysis of NAP in Southeast Asia reached the same conclusion (Chua et al. 2021).

Not all local stakeholders who will be affected by mandatory prescriptions in a few years’ time were aware of the regulatory changes, and most of the respondents at different levels did not believe that they would be implemented soon. In veterinary medicine in Vietnam, buying antibiotics over the counter is a common practice driven by farmers’ experience-based antibiotic usage (Bâtie et al. 2022; Carrique-Mas et al. 2015; Truong et al. 2019) as it is also in human medicine (McKinn et al. 2021). Moreover, in Vietnam, people without a practice certificate or veterinary training sell drugs (usually to a relative of the shop owner) and are therefore not allowed to issue prescriptions. In countries where over-the-counter drugs are highly regulated, the rate of self-medication is significantly lower than in Southeast Asia, where drugs are widely available. Self-medication leads to antibiotic misuse and overuse (Nepal and Bhatta 2018). Changing farmers’ habits (especially among the older generation and for small scale farms) was perceived as difficult in Vietnam because they are using antibiotics according to their own experience or their neighbor’s experiences. The implementation of this essential regulation in the fight against ABR will therefore require the development of tailored solutions.

### Solutions to improve the implementation of the new regulations

Several respondents stressed the need to clarify regulations and strengthen sanctions to improve their implementation. These sanctions should be more clearly defined, stricter, and systematic. This would require clear guidance, a distribution of tasks, and more resources for the authorities, including technical and human capacities (Jamrozik and Selgelid 2020). Participants suggested random on-farm feed inspections or targeting drug sellers to incur a domino effect. We also identified an informal flow of antibiotics leading to the sale and usage of banned substances. This informal flow has already been described in aquaculture systems (Brunton et al. 2019), and producers have reported using banned substances (Pham-Duc et al. 2019). Respondents also raised drug quality concerns, including the presence of diluted antibiotics, which led to higher antibiotic dosages or that expose the chickens to sub-therapeutic dosages. This finding was consistent with a previous study conducted among farmers in the Mekong Delta (Yen et al. 2019) and is also an issue in other Southeast Asia countries (Pham-Duc and Sriparamananthan 2021). This is a worrying situation as counterfeit or sub-standards antibiotics contribute to the emergence of ABR (Kelesidis and Falagas 2015).

Most respondents believe that reducing the proportion of small-scale farms in favor of large-scale farms and integrated systems would help to alleviate the ABR burden. Small-scale farms have often been blamed for antibiotic misuse and overuse in Vietnam and worldwide (Ducrot et al. 2021) and identified as stakeholders who fail to comply with regulations due to their lack of adaptability (Pham-Duc and Sriparamananthan 2021). The solution of reducing the number of small scale farms is in line with the government’s strategy outlined in the agriculture development plan for 2025 (Prime Minister 2021). This was also true in 2003, during the highly pathogenic avian influenza crisis, when industrial production was pushed through partnership between the public and agro-industrial sectors, because integrated farms were perceived as implementing better biosecurity (Figuié, Pham, and Moustier 2013). However, this study demonstrated that, due to consumer preference for the local market, only the most competitive ones developed. Also, as one respondent pointed out, increasing the size of farms does not always result in a reduction in ABU, and increasing size without adequate training can have negative consequences for ABU control. Moreover, intensive systems were also reported as heavy users of antibiotics, especially in prophylaxis (Bâtie et al. 2022; Luu et al. 2021). On the other hand, poor biosecurity in small-scall farms in Vietnam can also be challenging with outdoor systems and low farmers technicity (Thi Dien et al. 2023). Small-scale production has been shown in many regions to be a source of women’s empowerment, financial reserves, an important element for ceremonies, and a contributor to nutrition (Alders et al. 2018). It also represents social benefit and reinforces the relationship between householders (Dumas et al. 2018). Another study in Ethiopia found that as farm size increases, men usually take control, reducing the independence of women (Sambo et al. 2015). In Vietnam, the population is predominantly rural, and small-scale farmers still represent the most common production systems. Demand varies according to cultural events; for example, chickens are highly consumed during the Lunar New Year, and many smallholders only raise chickens for this occasion (Delabouglise et al. 2017). In Southeast Asia, the knowledge and practices of men and women regarding antibiotics differ depending on the context (education, socio-economic factors) (Pham-Duc and Sriparamananthan 2021). Thus, social factors must be taken into account when developing livestock production (Dumas et al. 2018; Sambo et al. 2015).

We found that small scale farms have difficulty complying with the new regulations because they lack the technical capacity to implement them. Indeed, respondents raised that small scale farms in Vietnam have poor farm management practice (biosecurity and density) as it has been reported in a study including key informants’ interviews (from governmental authorities, private sector, workers/producers, distributors research and educational institutions) (Thi Dien et al. 2023). Our respondents reported that poor management combined with the forecast conditions in Vietnam, contribute to higher disease incidence as it has already been shown elsewhere (Delabouglise et al. 2017; 2019). To compensate for the high diseases’ incidence, farmers use antibiotics in prevention to secure their production. Thus, respondents reported that farmers were afraid that banning this usage could lead to a loss of income. However, a survey of farmers, retailers, and veterinarians in Cambodia revealed that farms with higher levels of biosecurity used fewer antibiotics (Ornelas-Eusebio et al. 2020). Strengthening biosecurity is a target of the Vietnamese government, which is now developing biosecurity guidelines and projects focusing on biosecurity to improve the training of trainers with the National Agriculture Extension Center. Feed additives and probiotics are common in Vietnam and are necessary supplements, but respondents reported a lack of alternatives to antibiotics and the high price of these products.

In our study, the lack of knowledge among farmers and drug sellers, as well as some local authorities, was a barrier to the implementation of the new regulations. Because a lack of awareness of ABU and ABR’s broader impact drives ABU (Brunton et al. 2019), many studies have identified education programs as key interventions to improve ABU, which has increased the adoption of the regulations. Some respondents were aware that ABR can have serious consequences for human health, as highlighted in another study (Pham-Duc et al. 2019) and we found that reducing the ABR burden could also act as a lever in reducing ABU. Raising awareness already figures among the NAP objectives and has been carried over to the second NAP. In Vietnam, this is achieved through mass media channels, TV programs, or workshops organized at different levels. Respondents stated that, in addition to the communication campaign, more training from local stakeholders was required. Stakeholders must also be aware of the change in regulations, which did not seem to be the case for local stakeholders. According to the respondents, information on new regulations should be disseminated more widely, particularly locally. This could be done by organizing workshops with district or communal veterinarians. Finally, to raise compliance with regulations, relevant stakeholders must understand the objective of the law. Indeed, farmers did not always understand the link between ABU reduction and ABR, as well as the link between antibiotic residues and ABR, as observed in Cambodia (Chea et al. 2022). Farmers and drug sellers did not see the real benefit of the law, and as the fight against ABR is long-term, they tended to focus on their daily problems. In Cambodia, farmers were identified as focusing more on the benefits of their production rather than on the consequences of antibiotic use (Om and McLaws 2016).

Several stakeholders identified drugstores as critical components of the antibiotic reduction strategy. Moreover, according to the respondents, drugstores would be easier to manage than farmers because there were fewer of them. By raising awareness of good husbandry practices, they would then be able to provide farmers with advice. Mandatory prescription was sometimes perceived positively by this category of stakeholders as it would attract more customers and enhance the quality of advice. If farmers had fewer livestock diseases to manage, they would have higher incomes and would be willing to pay for the products. It would therefore be a win-win situation for them. In a previous study, drugstores were identified as the primary point of contact for farmers because they sell antibiotics and provide advice (Bâtie et al. 2022; Doan Hoang et al. 2019).

Increasing consumer willingness and showing the importance of producing better quality chicken could also have a leverage effect, as it has already been shown that consumers are afraid of chemical risks related to food safety (Nguyen-Viet et al. 2017). Producing safer food to meet consumer demand was also a motivation to comply with the regulations. Further studies are needed to assess the influence of consumers on changing the practices of veterinary drug value chain stakeholders.

## Conclusion

The development of a regulatory framework to reduce antibiotic misuse in chicken production in Vietnam is a necessary step towards changing ABU practices. However, this cannot be done in isolation and should be combined with other measures tailored to the specific context of implementation. This study identified levers that could be used to influence ABU practice.

Regulations in Vietnam were partially ignored because relevant stakeholders were too far away from the decision-making system and did not feel concerned about the regulations. There was also a lack of implementation due to insufficient resources (human, technical, and knowledge) among stakeholders in charge of enforcing the regulations, especially for small-scale farms. Improving communication at the local level on regulations and their purpose in the direction of local stakeholders would help to increase compliance. Moreover, new policies should consider the most vulnerable stakeholders, such as farmers and drug sellers.

## Acknowledgements

We are grateful to all the respondents who participated in the study. We are thankful to Nguyen Thi Nga for the data collection in north Vietnam and Le Thi Thu Hà and Ding Bao Truong for the data collection in south Vietnam. We would also like to thank Anita Saxena for the professional English proofreading. Part of this work has previously appeared online as part of the thesis of Chloé Bâtie.

## Data availability

De-identified data are available at: https://doi.org/10.18167/DVN1/9J57KF Specific requests must be submitted to the first author.

## Supplementary data

Supplementary data are available at: https://www.medrxiv.org/content/10.1101/2024.06.27.24309573v1.supplementary-material

Supplementary Material 1: guide for focus group discussion

Supplementary Material 2: guide for semi-structured interviews

Supplementary Material 3: classification of stakeholders

Supplementary Material 4: factors influencing the implementation of new regulations

