## Supplementary Material 1 for "Understanding the implementation of antimicrobial resistance policies in Vietnam: a multilayer analysis of the veterinary drug value chain"

Supplementary Material 1: Guide for focus group discussion of stakeholders of the veterinary drug value chain

### Guide – Focus group discussion

| **Stakeholder mapping**<br>Veterinary drug value chain in poultry production in Vietnam | |
|---|---|
| **Objectives:**<br>To draw the veterinary drug value chain with the identification of all stakeholders involved in it and to characterize interactions and roles of each stakeholder | |
| First step: identify all the stakeholders involved in the value chain | |
| Topics | Information required |
| - To identify all the stakeholders involved in the veterinary drug supply chain<br>➔ Post-it on the dashboard (different colors for private/public)<br><br>*Focus on Abs and then ask if the stakeholders are the same for the alternatives feed additives* | - Please, try to identify all the stakeholders involved in the Veterinary drug supply chain<br>- Public regulators (from central to local): MARD, DAH, DLP, …<br>- Research institution, university<br>- International organizations (research institutions, projects, cooperation, …)<br>- Private sectors (importers, pharmaceutical industries, feed industries, animal drugs sellers, integrators, farmers, …)<br>- Informal sellers |
| - What are the roles of each stakeholder in the value chain?<br><br>➔ Group all the stakeholders with the same roles on the dashboard | - Regulation<br>- Production/Importation<br>- Distribution<br>- Prescription<br>- Selling<br>- Usage<br>- Research/Project |
| Second step: draw the veterinary drug value chain | |
| Topics | Information required |
| - To draw the ABs flow in poultry production (including medicated feed)<br><br>➔ Draw the arrows of the flow of products<br><br>- To complete the map flow with proportion *(if possible, depending on the respondent)*<br><br>➔ Write the proportions on the arrows<br><br>*Draw the Abs flow and alternatives feed additives if different* | - Where the farmers can buy ABs for their poultry?<br>- Who are the sellers?<br>- How is organized the production/distribution/wholesale for ABs?<br>- Where do the ABs come from? *What is the proportion of imported products?*<br>- Are ABs sold abroad? *Where? What is the proportion?* Do you know of any illegal importation?<br>Do you know of any informal source of AB (middlemen, reselling….)? |

| | |
|---|---|
| - Identify the chain of advice<br><br>➔ Draw the arrow of the chain of advice<br>➔ The formality of the interaction: formal (ex with a contract), informal, both<br>➔ Strength of interactions | - What are the different users (different kinds of farmers can buy drugs from different sources)?<br>- When selling ABs, do they give advice?<br>- |
| - Identify the interactions between stakeholders<br><br>➔ Directions of interactions from one to another one, in both directions, network (include more than 2 stakeholders)<br>➔ The formality of the interaction: formal (ex with a contract), informal, both<br>➔ Strength of interaction | - What are the interactions or influences between the different stakeholders?<br>- Who works with whom?<br>- Who interacts with whom?<br>- What is the nature of the interactions?<br>- What is the link between the public and the private sector? (public-private partnership) |
| Third step: Identify the legislation | |
| - Identify the existing legislation | - What are the main legislations related to ABs that you are aware of related to Abs manufacture/sell/use?<br>- Are they applied? |
| - Identify the position regarding new regulations (if time)<br><br>*Ban of AB in the feed for prevention*<br>*Mandatory prescription* | - Are you aware of what is going to change in terms of regulations related to ABs?<br>- With this new regulation what is going to change in the interaction between the stakeholders? At which level? |
