## Supplementary Material 2 for "Understanding the implementation of antimicrobial resistance policies in Vietnam: a multilayer analysis of the veterinary drug value chain"

Supplementary Material 2: Guide for semi-structured interviews of stakeholders of the veterinary drug value chain

| **Questions** | **Action** |
| --- | --- |
| **Structural position and role in the value chain** | |
| Here is the value chain that has been drawn in a previous workshop.<br>Where do you stand on this value chain? | Presentation of the value chain on PADLET<br>Add on the diagram where is the stakeholders (5 stars on PADLET) |
| Could you please describe your activities?<br>For which company/institution are you working?<br><br>Could you please describe the activities of your company/institution?<br>Functions and missions of your institutions/companies in the command chain?<br><br>(How many people are in your company? When the company was established? What are the main products/services of your company? The volumes of products/services? ) | Complete the flow of antibiotic and/or alternative feed additives value chain<br><br>Role of the SH, of the company |
| **Interactions with other stakeholders** | |
| With whom are you working in your daily activities?<br>To whom do you sell your drugs? To whom do you buy the materials? Who are your clients? (think about exportation)<br><br>Who else do you interact with?<br>Could you please describe the nature of the interaction for each stakeholder?<br><br>Do your colleagues work/interact with other people? | Add to the diagram the additional stakeholders<br>Discuss for each interaction: the nature, direction, … by showing it on the diagram<br><br>Ranking on PADLET<br><br>Interactions |
| Are you a part of a professional organization for feed/drug companies/farm cooperatives?<br><br>Do you collaborate with the public sector or any international organization?<br>Do you collaborate with the private sector? (ie consultant for policy)<br><br>Which kind of collaborations? | Add on the diagram the collaborations |
| Do you trade/use/make policy on alternative medicines?<br>What is your opinion on these medicines?<br>Who are the main users? | Alternative feed additives |
| Who (authority) is performing the control? Do you have internal regulations decided by the company? | Add in the diagram the person in charge of the controls<br><br>Regulations |

| | |
|---|---|
| What kind of control is it? What is the frequency? | Interactions |
| **Knowledge and opinion on ABR and NAP** | |
| What is your opinion on ABR in Vietnam?<br>Do you know any strategies to fight ABR in Vietnam? | Opinion/Knowledge on ABR |
| *Since 2017, the Vietnamese government has implemented a strategy to fight against ABR in livestock production* | |
| Have you heard about it? If yes, from where?<br>What do you think about it? | Opinion/Knowledge on NAP |
| **Barriers / motivations new regulations** | |
| *Two new regulations have been issued: - ban on AB in feed for prophylaxis*<br>*- mandatory prescription to buy drugs* | |
| Were you aware of these two new regulations?<br>How do you know about the change in regulations? | Regulations<br>Knowledge on regulations |
| What do you think about those regulations?<br>Do you think that these changes is a good thing or a bad thing? For yourself and for the other stakeholders? | Interest |
| How do you think that these regulations are going to be applied? Why? | Willingness to implement the legislation |
| What are the positive and negative consequences? | |
| What do you think is going to change in the value chain?<br>What is going to change with these regulations in your daily activities? What will be the impact on your daily activities?<br>To compensate for these changes, what are you going to establish? | Change in the chain |
