## Supplementary Material 3 for "Understanding the implementation of antimicrobial resistance policies in Vietnam: a multilayer analysis of the veterinary drug value chain"

Supplementary Material 3: **Classification of the stakeholders of the veterinary drug value chain according to their level of legitimacy, resources, connections, and their role in the implementation of the new regulations on the progressive ban of AB in the feed (n°13/2020/ND-CP) and mandatory prescription (n°122020TT-BNNPTNT) from focus group discussion (n=12) semi-structured interviews (n=39) and analysis of the National Action Plan (NAP), 2021, Vietnam.**

| Stakeholder | Legitimacy | Resources | Connections | Role in the regulations |
|---|---|---|---|---|
| **Public sector** | | | | |
| *Government authorities* | | | | |
| Ministry of Agriculture and Rural Development (MARD) | NA | NA | NA | The MARD (Ministry of Agricultural and Rural Development) is divided into several institutions at the central level including among others the Department of Animal Health (DAH), the Department of Livestock Production (DLP), and the National Institute of Veterinary Research (NIVR). They are the final decision-makers in law enforcement. |
| Department of Animal Health (DAH) | +++ | ++ | ++ | Governmental authority at the national level, focal agency related to AMR issues (Veterinary Drug Management Division), control of importation and distribution of drugs and medicated feed, development of legal documents on veterinary prescription to buy drugs (n°122020TT-BNNPTNT), member of the Sub-National Steering Committee on ABR in the agricultural sector |
| Department of Livestock Production (DLP) | +++ | ++ | ++ | Governmental authority at the national level, control of importation and exportation of feed, control of feed companies together with DAH (for medicated feed), control of alternative feed additives, responsible for VietGAHP certificate, development of the legal document on the ban of AB in feed for prophylaxis (n°13/2020/ND-CP) |
| Sub-department of Animal Health and Livestock Production (Sub-DAHLP) | +++ | + | +++ | Governmental authorities at the provincial level, implement regulations, manage sales and use of antibiotics, provide and control practice certificates of veterinarians, control the distributors and stores selling antibiotics by samples testing (ban substances, authorization, drug quality), report veterinary activities to DAH, dissemination of information related to legislation to districts and communes, control of slaughterhouse (samples collection), license of domestic |

| | | | | |
|---|---|---|---|---|
| | | | | feed, test quality of the feed (samples collection), propose new regulations, guide farmers to implement VietGAHP standards |
| Provincial Department of Agriculture and Rural Development (DARD) | ++ | + | ++ | Governmental authorities at the province level, instruct agencies to manage and monitor the trading and use of AB, dissemination of legislation, provide guidance, checking and monitoring of the circular on prescription, provide training to prescribers, dissemination of information related to legislation to districts and communes, control of VietGAHP, issue VietGAHP certificate |
| Veterinary district station | + | - | + | Governmental veterinarians working at the district level under the SUbDAHLP, sometimes participate in the activities of SubDAHLP (control of stores that sell drugs and feed), management of the vaccination program, inspection of veterinary hygiene and food safety in a slaughterhouse, record demand to open stores to sell drugs in the district |
| Communal veterinarian | + | - | + | Communal veterinarians, mandated by the government, are responsible for the sanitary situation (reporting epidemics to the upper level), disease control, perform vaccination campaign |
| *National Research Centers* | | | | |
| National Institute of Veterinary Research (NIVR) | +++ | ++ | ++ | Provide evidence on ABU/ABR, member of the Sub-National Steering Committee on ABR in the agricultural sector, and recommendations to policymakers through FAO |
| University | NA | NA | NA | Provide evidence on ABU/ABR, member of the Sub-National Steering Committee on ABR in the agricultural sector, research on alternatives toward antibiotics, antimicrobial susceptibility laboratory testing, vocational training, and cooperation to improve professional skills |
| **Private sector** | | | | |
| *National private stakeholders* | | | | |
| Feed company | ++ | ++ | ++ | Sell feed, medicated feed (with veterinarian prescription), and provide technical advice to integrated farm and large family commercial farms |
| Alternative feed additives company | + | ++ | + | Importation, production, and sale of alternative products to feed company, agencies, or with technical advice to integrated farm and large family commercial farms |
| Importer | ++ | ++ | ++ | Record importation and sale of AB, technical advice to integrated farms and large-scale family commercial farms, feedback on law proposals |

| | | | | |
|---|---|---|---|---|
| Producer | ++ | ++ | ++ | Record purchase and use of AB materials, feedback on laws proposal, technical advice to integrated/large scale family commercial farms |
| Distributor | ++ | + | ++ | Record importation and sale of AB to SubDAHLP, sell AB with prescription, feedback on laws proposal, technical advice to integrated/large scale family commercial farms, have to register to the DAH |
| Technician | + | ++ | ++ | Provide AB and technical advice to integrated farms |
| *Local private stakeholders* | | | | |
| Agency level 1 | + | + | ++ | Record importation and use of AB, sell AB that is on the list of permitted drugs for circulation, technical advice to farms, autopsy, sample collection, farm visit, need certification to trade drugs/business license (every 5 years), need to have at least a veterinary intermediate training degree |
| Agency level 2 | + | + | + | Sell AB, and technical advice to household and small family commercial farms, autopsy, sample collection, farm visit |
| Veterinarian | + | + | + | Sell and administer AB, farm visit, autopsy, technical advice, clinal examination |
| *Informal value chain* | | | | |
| Informal drug seller | NA | NA | NA | Sell AB without a practicing certificate or certificate to sell |
| Human pharmacy | NA | NA | NA | Sell AB for animals in human pharmacies |
| *Users* | | | | |
| Integrator | NA | NA | NA | Chicken company that can also own a drug and or feed plant as well as farms (the others are under contract) and slaughterhouses. They perform controls at the farm level and in the slaughterhouses and provide technical support to the farmers as well as all inputs (feed, DOCs, drugs) and collect the chickens at the end of the production cycle, feedback on the law proposal |
| Integrated farm | + | ++ | ++ | More than 2000 chickens, intensive system in contract with a chicken company or integrated to the company, confined system, use AB for treatment and prevention according to the instruction of the technician hired by the company, produce mostly exotic chickens (white chickens) |
| Family commercial farm | + | + | ++ | From 100 chickens to thousands, predominance of the semi-confined system, source of AB varies according to the size, use AB for treatment and prevention according to drugstores' instruction or from their own experience, produce mostly hybrid chickens (or colored chickens) |

| | | | | |
|---|---|---|---|---|
| Household farm | + | + | + | Small producers, less than 100 chickens, free-range, mainly for self-consumption, small supply, use AB for treatment, produce mostly local breed chickens |
| VietGAHP farm | NA | NA | NA | Farms with a VietGAHP certificate (voluntary program), must comply with requirements (record ABU, no AB residues, located outside residential area, …), allowed to sell to supermarkets |
| State farm | NA | NA | NA | |
| **International partners** | | | | |
| World Health Organisation (WHO) | NA | NA | NA | Provide guidelines on good ABU practice and ABR, push for the development of the NAP |
| Food and Agriculture Organisation (FAO) | ++ | +++ | ++ | Provide evidence on ABU/ABR, contribution on development of the NAP, review on the legislation, support of the government |
| World Animal Health Organisation (WAHO) | NA | NA | NA | Provide guidelines on good ABU practice and ABR, push for the development of the NAP, DAH report ABU to them every year |
| International research center | ++ | ++ | ++ | Provide evidence on ABU/ABR, contribution on the development of the NAP, recommendations to policy-makers |
| International collaboration | ++ | +++ | + | Technical expertise on law design on the use of AB in the feed and prescription, technical assistance project |
| Donor | NA | NA | NA | Financial support on research project on ABU/ABR and development of the NAP |

**Legitimacy**: Defined by the institutional position (acquired by law or perceived by the public to be legitimate) of the stakeholders and their involvement in the law design and/or by the type of antibiotics flow formal (flow monitored by the government) or informal. **Resources**: Defined by stakeholders' knowledge of ABU, ABR, and regulations, and by the technical (financial, human, material) resources to apply the new regulations. **Connections**: Number of interactions within the veterinary drug value chain and quality of stakeholder's relationships (level of trust, frequency of connection, formal or informal). +++ strong, ++ medium, + weak, – absent, ? indetermined. NA: not assessed. Definitions adapted from Zimmermann, 2007; Poupaud et al., 2021; Bordier et al.,2018. (19–21
