## Supplementary Material 4 for "Understanding the implementation of antimicrobial resistance policies in Vietnam: a multilayer analysis of the veterinary drug value chain"

Supplementary Material 4: **Factors influencing the implementation of the new regulations to reduce antibiotic use in chicken production in Vietnam from respondent's perspectives from semi-structured interviews (n=39) 2021, Vietnam.** MARD: Ministry of Agriculture and Rural Development; DAH: Department of Animal Health; DLP: Department of Livestock Production; SubDAHLP: Subdepartment of Animal Health and Livestock Production; DARD: Department of Agriculture and Rural Development;

| **Stakeholder** | **Main barriers** | | | | | | **Main enablers** | | | |
|---|---|---|---|---|---|---|---|---|---|---|
| | Level of knowledge | Technical guidance | Conflict of economic interest | Scale-related management discrepancies | Technical and financial capacities | Informal distribution channels | Stakeholder's perceptions | International influence | Consumers demand for food safety | Reducing the burden of ABR |
| **Public sector** | | | | | | | | | | |
| *Government authorities* | | | | | | | | | | |
| DAH/DLP | | | | | | | | | | |
| SubDAHLP | | | | | | | | | | |
| DARD | | | | | | | | | | |
| Veterinary district station | | | | | | | | | | |
| Communal veterinarian | | | | | | | | | | |
| *National Research Centers* | | | | | | | | | | |
| **Private sector** | | | | | | | | | | |
| *National private actors* | | | | | | | | | | |
| Feed company | | | | | | | | | | |
| Alternative feed additives company | | | | | | | | | | |
| Importer | | | | | | | | | | |
| Producer/Distributor | | | | | | | | | | |
| Technician company | | | | | | | | | | |
| *Local private actors* | | | | | | | | | | |
| Agency level 1 and 2 | | | | | | | | | | |
| Private veterinarian | | | | | | | | | | |
| *Users* | | | | | | | | | | |
| Integrated farm | | | | | | | | | | |
| Family commercial farm | | | | | | | | | | |
| Household | | | | | | | | | | |
| **International partners** | | | | | | | | | | |
